# Timing and nature of palliative care discussions are patient-specific according to clinicians: a qualitative study

**DOI:** 10.1101/2019.12.28.19013417

**Authors:** N Tavares, KJ Hunt, N Jarrett, T M A Wilkinson

**Affiliations:** Solent University; NIHR CLAHRC Wessex; University of Southampton; University of Portsmouth; University Hospital Southampton

**Author notes:** Corresponding author’s contact details: Nightingale Building, Faculty of Health Sciences, University Road, Southampton SO17 1BJ.

**Keywords:** COPD, palliative care, communication, patient-clinician communication, patient-centre care, qualitative research

## Abstract

**Background:** Chronic obstructive pulmonary disease (COPD) is associated with an unpredictable and complex disease trajectory. Consequently, most patients are not involved in advance care planning and do not receive palliative care until the end of life.

**Aim:** To explore clinicians’ experiences, opinions and recommendations for the timing and nature of palliative care discussions in COPD.

**Design:** Qualitative interviews with nurses and doctors that provide direct care to COPD patients.

**Setting/Participants:** 14 clinicians working across primary and secondary care in the UK were interviewed.

**Results:** Participants suggested that those with the appropriate expertize and an established relationship with patients were best placed to initiate discussions about palliative and future care. Early, gradual and informed palliative and future care discussions were considered best practice, however they uncommon occurrence due to service, patient and clinician-related barriers. The unpredictable disease trajectory and fine balance between providing acute care and discussing palliative care options were suggested as key greatest barriers for discussions. However, damaging patient hope was a concern for clinicians and reduced their inclination to discuss palliative care. Clinicians did not seem to think that patients were ready for discussions, therefore they avoided broaching the subject leading early in the disease trajectory.

**Conclusion:** Stand-alone conversations about and near the end of life was described as current usual practice by clinicians, however individualised early, regular and gradual discussions with patients about immediate and long-term future plans may make such discussions feel less negative and ordinary for patients and clinicians.

## Background

Chronic obstructive pulmonary disease (COPD) is a progressive and life-limiting illness that causes breathlessness and chronic cough [1, 2]. COPD patients have a high symptom burden [3, 4] and these symptoms are managed by means of aggressive and invasive treatments, especially as they approach the end of life [5]. Palliative care can support these patients holistically and provide care that focuses on quality of life and effective symptom relief [6]. In order to offer care focused on patients’ preferences, conversations about future care need to take place [7, 8]. However, the unpredictability and complexity of COPD prevents the early identification of patients in need of palliative care [8, 9]. This means that there is often no obvious ‘right time’ to initiate conversations about end of life and palliative care.

Discussing preferences and creating end of life care plans can ultimately result in improved experiences at the end of life while reducing patients’ symptom burden [10, 11]. Despite this, most COPD patients do not get the chance to discuss their preferences until they approach the end of life [12, 13]. A recent paper has focused on understanding COPD patients’ preferences for palliative and future care discussions with patients [14]. Clinicians’ opinions about COPD patient preferences for discussions are currently unknown [12], so this study aimed to explore clinicians’ perspectives on when and how to initiate and conduct palliative care discussions with COPD patients.

## Methods

### Design

This paper reports on the third component of a larger study looking at how and when to start conversations about palliative and end of life care planning in COPD. A systematic literature review [12] and a qualitative study of patients’ preferences and opinions for the timing and nature of discussions [14] formed the first components of the study. These two components informed the design of this study, the recruitment strategy and the interview topic guide (such as patient perceptions about the lack of expertise in primary care and their preference in delaying these discussions). Recruitment focused on health professions cited by patients as the preferred person to discuss palliative care [14].

### Sample

The study used a purposive sample and recruited clinicians who provided direct care to COPD patients at different stages of their condition. This included practice nurses; general practitioners (GPs); COPD consultants and COPD specialist nurses. It was postulated that these clinicians worked in environments where discussions could be held sensitively; and had some training or knowledge about palliative care in COPD. The inclusion criteria can be found in **Table 1**.

**Table 1.** Inclusion criteria for healthcare professionals

| **Table 1 Inclusion criteria for healthcare professionals** |  |
| --- | --- |
| **Professional background** | Practice nurses, GPs, COPD consultants and COPD specialist nurses |
| **Age** | Aged 18 or over |
| **Clinical practice** | Clinicians providing direct care to COPD patients |
| **Language** | English speaking clinicians |
| **Location** | Clinicians working in geographical area of Hampshire |

Clinicians were recruited by email or by letter. The NIHR Clinical Research Network (CRN) signposted primary care clinicians to the study. The CRN informed GP practices about the research study and requested that clinicians willing to participate contacted the first author. Secondary care clinicians were signposted to the study by the research department of the hospital where they worked. Clinicians were sent information about the study and were requested to contact the researcher if willing to participate. Written informed consent was obtained for all participants.

### Data collection

Interviews were audiotaped, lasted 20-45 minutes and followed a semi-structured approach. The interviews were conducted by the first author (NT^1^), who had no previous relationship with the participants. The questions followed a topic guide that focused on exploring clinicians’ understanding about the nature and timing of palliative care discussions and in debating patients’ preferences for these discussions. The exact phrasing of the questions and the scope and depth of the interview varied on an individual basis.

Individual interviews were undertaken at a time and place convenient to the participant, usually GP practices and hospital. Data collection was carried out between December 2018 and March 2019 and stopped at data saturation point. Data saturation was judged achieved when no new themes/topics were emerging from the interviews with clinicians. The decision to stop data collection was discussed and agreed within the authorial team.

### Data analysis

All interviews were transcribed verbatim and anonymised. Data was managed using qualitative software Nvivo 11 and analysed using thematic analysis [15, 16]. Transcripts were read several times and coded. Codes were developed, defined, reviewed and grouped into larger categories of codes and final themes. All transcripts were analysed by the first author (NT), whilst all the authors monitored and supervised the transcript analysis, participated in the development of the codes and themes, contributing to the trustworthiness and methodological quality of the study. The consolidated criteria for reporting qualitative research (COREQ) were used to report the research findings [17].

## Results

Fourteen clinicians, out of 22 clinicians contacted, participated in the study and their characteristics are summarised in **Table 2**.

**Table 2.** Clinicians’ characteristics

| Table 2 Clinicians' characteristics |  |
| --- | --- |
| Gender – Male (%) | 4 (29%) |
| Mean age in years (SD) | 50 (8.5) |
| Years in practice (SD) | 22 (12.5) |
| Number of COPD patients seen a week (SD) | 17 (8) |
| Number of discussions a monthly (SD) | 3 (5) |
| Special interest or training palliative care (%) | 6 (43%) |
| Professional background: |  |
| • COPD consultants | 3 (23%) |
| • COPD specialist nurses | 3 (23%) |
| • Practice nurses | 4 (27%) |
| • General practitioners | 4 (27%) |

The analyses yielded 59 deductive codes and 50 inductive codes. The deductive codes focused on addressing the aim of the study, whereas inductive codes provided context and further information on clinicians’ thoughts. The combination of the different codes originated 7 categories of which 4 themes emerged, please refer to **Table 3**.

**Table 3.**
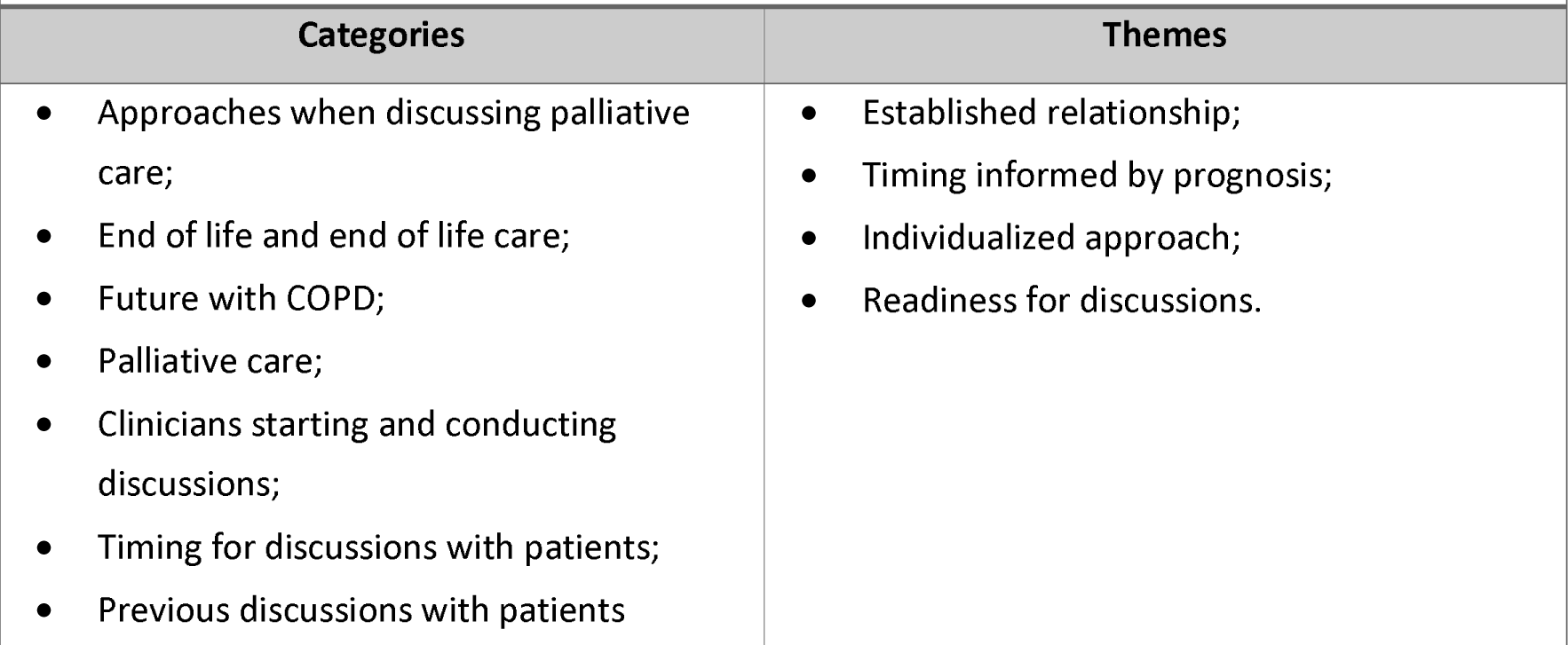
Categories and themes generated from interviews with clinicians

|  |  |
| --- | --- |
| Categories | Themes |
| • Approaches when discussing palliative care;<br>• End of life and end of life care;<br>• Future with COPD;<br>• Palliative care;<br>• Clinicians starting and conducting discussions;<br>• Timing for discussions with patients;<br>• Previous discussions with patients | • Established relationship;<br>• Timing informed by prognosis;<br>• Individualized approach;<br>• Readiness for discussions. |

Quotes were selected purposively and unnecessary information or irrelevant pauses were edited out for clarity and anonymity.

### Established relationship and clinician expertise

Healthcare professionals could not pinpoint specific clinicians that should have palliative care discussions with patients. Instead, most clinicians believed that patients should discuss this with a well-known clinician, since this was thought to increase trust and familiarity. Participants believed that some clinicians felt less comfortable when discussing palliative care with patients, since some patients became emotionally distressed. Despite this, no participant found discussions particularly difficult for themselves.

When analysing interdisciplinary differences, more medics considered themselves suitable clinicians to initiate discussion than nurses. Some primary care clinicians believed that hospital clinicians should take ownership of discussions, since care plans developed in primary care were not always respected in hospital. Most participants believed that communication skills and knowledge about COPD, palliative care and services available were essential. Participants’ knowledge and skills differed across and within professional backgrounds. However, most participants seemed to have little understanding about palliative care, since they viewed it as end of life care and exclusive of acute treatments.

### Secondary care

Consultants reported regular palliative care discussions with patients in outpatient clinics, especially on hospital wards. Participants believed that consultants’ medical background; expertise; patient-relationship; and regular contact with end-stage patients made them suitable clinicians to start discussions with patients. COPD specialist nurses had daily discussions with patients and provided palliative care to patients in the community. Moreover, these clinicians seemed to have a large knowledge base, the opportunity to build relationships with patients, and offer longer appointments. However, these clinicians did not always have the confidence to initiate discussions.

> *I’ve just been doing that (patient support sessions) depending on the patients. But it definitely gives me a better amount of time and relationship with patients. (…) I think in consultant clinics you get to develop a good relationship with patients if you do the same clinic, which I do. Because you get a caseload of patients and you get to know them*.

### Primary care

Participants stated that increasingly higher pressure in primary care resulted in shorter appointments, longer waits and reduced continuity of care. In fact, one practice nurse highlighted the variation in GP surgery management, which meant that patients were left in a *“postcode lottery”* for care. Some nurses and GPs were *“thrown in”* to COPD clinics with little to no training, so clinician skill and competency varied greatly across surgeries. Since primary care clinicians managed patients with different health conditions, some participants believed they possessed a less in-depth knowledge about COPD.

> *You can get one practice where the practice nurses are specialists in their own rights. Because they’ve done courses, they’ve done the spirometry course and all of that, and a COPD course. But then there are COPD nurses out there that have done very little courses that pick it up as they go along. (P9, Practice nurse)*

GPs felt that their overview of the patient’s medical history provided them with the required tools to start discussions. However, in contrast to practice nurses, GPs felt less able to build relationships with patients and had less time for discussions. Despite this, most practice nurses did not discuss palliative care, since they lacked the necessary training and knowledge. Moreover, practice nurses only saw patients with milder COPD, who nurses believed did not require palliative care discussions.

> *I mean it’s harder for me now (to build relationships with patients), because I’m only doing one day a week here. I don’t think it’s difficult (to build relationships). I mean I think things have changed in general practice. In that we don’t see those patients as often, as I say the practice nurses do the respiratory reviews and stuff. So they are more likely to have those conversations. (P12, GP*

### Timing informed by prognosis

Clinicians had conflicting thoughts about the ideal timing for discussions with patients. Timing was based on the individual patient and on poor prognosis. Participants found prognostication complex, so looked for factors that signalled poor prognosis when timing discussions, such starting long-term oxygen. In general, participants started discussions with patients with an advanced and deteriorating condition, usually at the end of life.

> *You can be now very severe (COPD), end stage (COPD), but that end stage could be weeks, months, could be years. We don’t tackle or approach it (palliative care discussions) with patients, because the trajectory is so unknown and varied. You could be saying we’re going to do this end of life stuff, but they might still be here in 18 months. We’ve got loads of them (patients) that you’re surprised they’re still going, but I wouldn’t be surprised if somebody said “they died”. I’d be like “okay it’s sad, but not unexpected*.*”*

> *(P4, COPD nurse)*

In contrast to current practice, most participants believed all patients should be given the opportunity to start early discussions and that early discussions could reduce the emotional impact of a late discussion, allowing patients to plan ahead. In fact, due to the complex nature of COPD, one consultant believed that care should steadily transition from aggressive to more comfortable treatments/care as disease severity progressed. Moreover, participants recommended regular discussions with digestible information that increased in depth as the patient’s condition deteriorated. Annual reviews and pulmonary rehabilitation were considered ideal opportunities to start discussions and offer education about palliative care.

> *If it’s someone I know well, that’s something (palliative care) I like to broach early on if I can. If it’s someone you can see declining just over a period of time. At least having it mentioned once or twice, then when you really want to discuss it, it doesn’t feel unusual to the patient. It feels like it was something you were naturally coming to*.

### Individualized approach

Although, participants reported that both parties started the discussion, they started discussions much more often than patients. Participants highlighted that patients and families rarely started discussions in a direct way, instead they provided cues about their readiness to start discussions. The lack of discussions started by patients was attributed to their poor understanding about the severity and progressive nature of their condition, and in differentiating between exacerbations and chronic decline.

> *I usually start it (the discussion), very rarely you get a patient (to start it). Actually, I suppose I’ve had one or two that talked about it in a roundabout way, because a family member had died of COPD and that sometimes leads to conversations at the start, “I don’t want to be like that person, because they were so breathless at the end*.*” But it’s a minority. (P3, Respiratory consultant)*

Many factors influenced the approach chosen by clinicians when starting discussions with patients, including their anxiety levels and understanding about their disease severity. Participants’ approach to starting and conducting discussions varied between direct and indirect. These approaches can be found in **Table 4**.

**Table 4.**
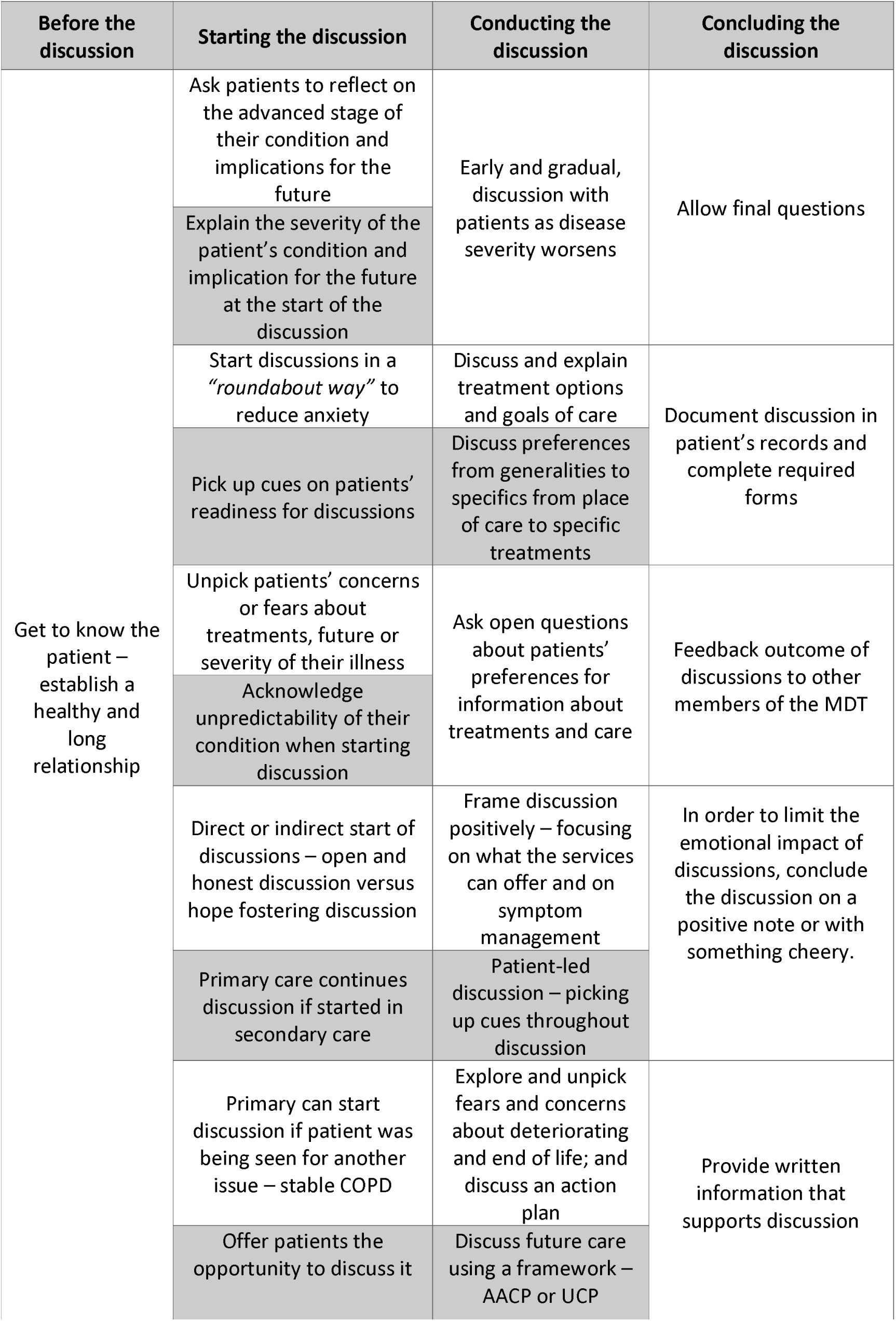
Methods used/recommended when discussing palliative care with patients

| Before the discussion | Starting the discussion | Conducting the discussion | Concluding the discussion |
| --- | --- | --- | --- |
| Get to know the patient – establish a healthy and long relationship | Ask patients to reflect on the advanced stage of their condition and implications for the future | Early and gradual, discussion with patients as disease severity worsens | Allow final questions |
|  | Explain the severity of the patient's condition and implication for the future at the start of the discussion |  |  |
|  | Start discussions in a *"roundabout way"* to reduce anxiety | Discuss and explain treatment options and goals of care | Document discussion in patient's records and complete required forms |
|  | Pick up cues on patients' readiness for discussions | Discuss preferences from generalities to specifics from place of care to specific treatments |  |
|  | Unpick patients' concerns or fears about treatments, future or severity of their illness | Ask open questions about patients' preferences for information about treatments and care | Feedback outcome of discussions to other members of the MDT |
|  | Acknowledge unpredictability of their condition when starting discussion |  |  |
|  | Direct or indirect start of discussions – open and honest discussion versus hope fostering discussion | Frame discussion positively – focusing on what the services can offer and on symptom management | In order to limit the emotional impact of discussions, conclude the discussion on a positive note or with something cheery. |
|  | Primary care continues discussion if started in secondary care | Patient-led discussion – picking up cues throughout discussion |  |
|  | Primary can start discussion if patient was being seen for another issue – stable COPD | Explore and unpick fears and concerns about deteriorating and end of life; and discuss an action plan | Provide written information that supports discussion |
|  | Offer patients the opportunity to discuss it | Discuss future care using a framework – AACP or UCP |  |

Some participants felt that starting discussions in a *“roundabout way”* (indirect approach) reduced anxiety levels and made discussions more acceptable. This approach was especially used in patients with increased anxiety and depression levels. The use of strong words, such as “palliative care,” “death” and “end of life,” were avoided, since these were seen to cause distress and reduced hope. In contrast, a small proportion of participants believed they should have an open and honest discussion with patients (direct approach), by providing accurate and direct information about their advanced condition. Despite the approach chosen, most discussions focused on treatment withdrawal.

> *I don’t think I would be talking about palliative care with them. I would be talking to them as an individual and how they feel about what’s going to happen? And how do they feel about dying? I wouldn’t probably mention the word palliative, because they wouldn’t understand what that means. But it might come when you’re talking about introducing the palliative care team. (P9, Practice nurse)*

## Readiness for discussions

Participants looked for prompts that could help them identify when patients were ready to initiate discussions. They suggested that patient readiness to discuss palliative care depended on various factors (**Table 5**). However, participants’ perception of patients’ readiness also seemed to impact their own readiness to initiate discussions.

**Table 5.** Factors associated with readiness for discussions

| Table 5 Factors associated with readiness for discussions |
| --- |
| • Patient’s older age;<br>• Patient’s understanding about COPD and current status of their condition;<br>• Patient’s perception about palliative and future care discussions;<br>• Patient’s attitude towards death;<br>• Patient’s personality;<br>• Patient’s or other people’s experiences regarding healthcare;<br>• Clinicians’ approach when starting and conducting discussions;<br>• Focus of discussion. |

Clinicians believed that patients’ understanding about the progressive nature of COPD and their disease severity impacted patients’ readiness to discuss palliative care and was related to insufficient information provided to patients throughout their disease trajectory. In fact, some participants had purposely withheld information about the progressive nature of COPD, for fear that this would emotionally impact patients. However, providing education about the progressive character of COPD was seen as very important.

> *I did (talk to patients about palliative care) when they were first diagnosed in a sense. Because I talked to them about what COPD was and I’d give them the booklet. I tried to avoid it being an aggressive or progressive disease. I didn’t say anything about palliative care. I would say if you carry on smoking things might worsen, you might end up on oxygen and things like that. (P9, practice nurse)*

## Discussion

### Summary of the findings

This study explored clinicians’ experiences of discussing palliative care with patients. Clinicians felt that those with a relationship with the patient should start the discussion. Discussions were started when the end of life was identified, through the recognition of key clinical red flags. In contrast to current practice, early, gradual and regular discussions with patients were seen as best practice. Direct or indirect approaches were used when starting and conducting discussions. Whether discussions came late or early in the relationship or took a direct or indirect approach, the discussion was tempered by the need to maintain hope and a good relationship, but also to avoid raising false expectations on what resources were available for patients and what medicine could achieve for them.

### Discussion of findings

Clinicians believed that discussions should ideally be started by clinicians with increased expertise and good relationships with patients. Therefore, there was no agreement on which clinicians were the most suitable to discuss this. Previous research has also highlighted a lack of clarity on who’s responsible for initiating discussing with patients [10]. Although clinical expertise and patient-relationships are paramount when discussing palliative care [18, 19], the lack of a designated clinician responsible for initiating discussions with patients can lead to nobody starting them. However, there seems to be a general lack of research exploring the lack of an explicit clinician responsible for discussing palliative care.

The absence of a designated clinician who is responsible to start palliative care conversations with patients may result in discussions not being started or being poorly conducted, due to lack of training and experience on conducting these discussions. Previous research suggested that clinicians with less expertise are less likely to start and conduct palliative are discussions, since expertise is dependent on training and in discussions taking place on a regular basis [19]. Specific guidance on who is responsible to start discussion, and on the timing and approach for palliative care conversations may facilitate the development and implementation of interventions that can help improve the frequency and quality of discussions [10]. Thus, guidance may reduce clinicians’ uncertainty regarding the timing and nature of discussions, increase clinicians’ confidence, which could result in increased willingness and reduced emotional impact of discussions on patients [10].

A recurrent theme from interviews was the challenges faced by primary care. The increasingly large workload and staff shortages has led to a reduction in the amount and quality of care provided to patients [20, 21]. Short appointments have also been highlighted in previous studies [22, 23], which have led to a reduction in direct patient care [24]. GPs found it particularly difficult to build patient-relationships and to find time for discussions, whilst practice nurses lacked the time and knowledge. As a consequence, the barriers found in primary care prevented the integration of early palliative care discussions with patients [25]. As a result, patients had to wait until they engaged with secondary care and their condition deteriorated considerably, before having the opportunity to start discussions. The integration of discussions could start while a patient’s condition is stable and shortly after diagnosis [25]. Primary care clinicians felt able to start early conversations if time and training was provided.

Throughout the interviews there was conflicting opinions about best and current practice for the timing of discussions. Most participants had a one-off discussion with patients usually at the end of life, whilst they believed discussions should be started earlier and be held regularly with patients. Late or lack of discussions have also been reported in other life-limiting conditions, such as heart failure and chronic kidney disease [26, 27]. Starting discussions late in the disease trajectory seems to be related to clinicians’ poor understanding of palliative care (confined to the end of life) and to the difficulty of prognosticating in COPD [28-30]. As an example, the practice nurses interviewed reported a lack of training and confidence to start discussions. Despite this, a previous study showed that a proportion of patients would like to discuss palliative care with practice nurses [14]. These differences in patients’ and clinicians’ perceptions about the timing and nature of palliative care seem to seriously impact their occurrence. Service rationing, clinician and patient-specific barriers, such as unpredictable disease trajectory and lack of time [28, 31], were also barriers for early discussions with patients. A change in the current model of care, from stand-alone discussions at the end of life, to early, gradual and brief discussions throughout the disease trajectory could minimize these barriers [25, 32].

Interestingly, the importance of social determinants was seldom mentioned by clinicians, despite being commonly reported in previous research [33, 34]. Low socioeconomic status can negatively impact patients’ understanding and perception of their condition, reduce their ability to self-manage, and decrease their ability and willingness to discuss palliative care [33, 35, 36]. Further, low socioeconomic status were associated with reduced knowledge and power, which are important factors when accessing and negotiating care [34, 35, 37]. Lack of power and knowledge can lead patients to devolve all expertise and decision power to clinicians, which limits their participation in palliative care discussions [38, 39]. Poor understanding and low health literacy seems to be related to a lack of information provided by clinicians about patients’ condition, especially about their progress over time [10, 14, 40]. In order to improve access to healthcare, and palliative care in particular, patients need education, through early and informative discussions about all aspects of COPD, palliative care and future treatments. Late discussions further reduce patients’ power, since they will often be at their most vulnerable at the point at which they require palliative care.

## Limitations

The first limitation of the study is that most clinicians expressed some interest in palliative care discussions. This was not the case for two nurses – one practice nurse and one COPD specialist nurse. Clinicians interested in the topic may feel more confident and comfortable in discussing palliative care. Interviewing clinicians less interested in discussing palliative care or that found discussions particularly difficult, could have generated different data. Another limitation of the study is the lack of ethnic diversity in the sample which prevented any exploration of the impact of ethnicity on palliative care discussions. This may have been especially important when considering the spiritually attached to palliative care discussions.

## Conclusion

Clinicians believed that a person with increased expertise and an established relationship with the patient should start palliative care discussions. Various clinician-specific barriers mean that there is no designated clinician that should start discussions. Current stand-alone discussions are difficult to time and to conduct appropriately and sensitively, resulting in lack or late discussions with patients.

Instead, discussions should be started from diagnosis, develop over time and information should be provided gradually. The discussion should be patient-led and patient-centred, focused on patients’ preferences for immediate care and held in a comfortable and private environment. A shift in the model of care, from stand-alone end of life care conversations to integrating palliative care early in the disease trajectory, is required.

## Data Availability

Data may be obtained from a third party and are not publicly available

## Authorship declaration

All authors participated in the different components of the reaserch study.

## Research Ethics

The study was approved by the Health Research Authority and the Hampshire B Research Ethics Committee in February 2017 with the following IRAS ID number: 203444.

## Data management

All data are kept in the University of Southampton repositories for a minimum of 10 years. For more information please contact the University’s Data Protection Officer at.

## Acknowledgments

The authors are grateful to all clinicians that took their time to participate in the research study.

## Footnotes

1 Clinical Doctoral Research Fellow – Staff Nurse and PhD Student. Training completed on qualitative interviewing

